# A quantitative exploration of symptoms in COVID-19 patients: an observational cohort study

**DOI:** 10.1101/2020.10.26.20220137

**Authors:** Gaojing Qu, Junwen Chen, Guoxin Huang, Meiling Zhang, Hui Yu, Haoming Zhu, Lei Chen, Dengru Wang, Bin Pei

**Author notes:** **Correspondence to:** Bin Pei, Evidence-Based Medicine Center, Xiangyang No.1 People’s Hospital, Hubei University of Medicine, 15 Jiefang Road, Fancheng District, Xiangyang 441000, Hubei, China., Dengru Wang, Yunnan Yanling Biological Technology co., LTD, Aerospace and IT Technology Park, 1389 Changyuan North Road, Gaoxin District, Kunming 650224, Yunnan, China. Bin Pei and Dengru Wang contributed equally to this article. **First author:** Gaojing Qu and Junwen Chen contributed equally to this article.

## Abstract

**Background:** As the spreading of the COVID-19 around the global, we investigated the characteristics and changes of symptoms in COVID-19 patients.

**Methods:** This was an ambispective observational cohort study, and 133 confirmed COVID-19 patients were included and all symptoms over the course were analyzed qualitatively. The symptoms, their changes over the course in the cohort and in the different clinical types, etc. were illustrated. Differences in different periods and severities were analyzed through Chi square test, association with severity was analyzed through LASSO binomial logistic regression analysis. Inter-correlation and classification of symptoms were completed. Major symptoms were screened and their changes were illustrated.

**Results:** A total of 43 symptoms with frequencies as 6067 in this cohort. Differences of symptoms in different stages and clinical types were significant. Expectoration, shortness of breath, dyspnea, diarrhea, poor appetite were positively but vomiting, waist discomfort, pharyngeal discomfort, acid reflux were negatively correlated with the combined-severe and critical type; dyspnea was correlated with the critical type. The 17 major symptoms were identified. The average daily frequency of symptoms per case was decreased continuously before the transition into the severe type and increased immediately one day before the transition and then decreased. It was decreased continuously before the transition date of the critical type and increased from the transition into the critical type to the next day and decreased thereafter. Dyspnea (*P*<0.001), shortness of breath (*P*<0.01) and chest distress (*P*<0.05) were correlated with death and their corresponding coefficient was 0.393, 0.258, 0.214, respectively.

**Conclusion:** The symptoms of COVID-19 patients mainly related to upper respiratory tract infection, cardiopulmonary function, and digestive system. The mild type and the early stage in other types mainly related to upper respiratory tract infection. The cardiopulmonary function and digestive system associated symptoms were found in all other types and stages. Dyspnea was correlated with critical type and dyspnea, shortness of breath, and chest distress were correlated with death. Respiratory dysfunction (or incompleteness) associated symptoms were the characteristic symptoms. The changes of symptoms did not synchronously with the changes of severity before the transition into the severe or critical type.

## Introduction

The Corona Virus Disease 2019 (COVID-19), caused by SARS-CoV-2 infection ^[1-4]^, has spread around the world and become a global pandemic declared by World Health Organization since March 11, 2020 ^[5, 6]^. Currently, there is no specific vaccine and antiviral drugs for COVID-19. Precise diagnosis and subsequent proper treatment according to the course was important.

Currently, the diagnosis of COVID-19 mainly relies on symptoms, laboratory tests, and chest computerized tomography. Symptom, a indicator reflecting discomfort of patients, was one of the most important clinical manifestations, although it was subjective and affected by self-sensitivity and expressing willingness. The variety and frequency of symptom can reflect the severity of the diseases ^[7]^.

Studies including Zhou *et al*. and Chen *et al*. have revealed that most COVID-19 patients exhibit fever. Cough, expectoration, and myalgia are also commonly symptoms in the COVID-19 patients. Dyspnea was also reported in COVID-19 confirmed cases ^[8, 9]^. Additional studies have showed that COVID-19 may cause symptoms related to the nervous system ^[10]^, cardiovascular system^[11-13]^, urinary system ^[14]^, skin ^[15,16]^, taste and smell ^[17]^. Li *et al*. disclosed the occurrence rate of fever, cough, expectoration, headache, and other symptoms in COVID-19 cases from Zhejiang, China. The results suggested that the cases in Zhejiang were mainly mild and moderate cases, which was significantly different from Wuhan ^[18]^. However, studies on symptoms of COVID-19 are only descriptive now ^[19, 20]^. With the global spread of COVID-19, more comprehensive and systematic studies were needed.

The purpose of this study was to investigate the development and clinical characteristics based on symptoms in a cohort with 133 COVID-19 patients. In this study, we conducted a quantitative analysis on the symptoms in frequency and occurrence rate, analyzed the association with different clinical types, and classified the symptoms based on pathophysiology, etc. to provide theoretical foundation for the prevention and diagnosis of COVID-19.

## 1. Materials and Methods

### 1.1 Design

This study is an ambispective observational cohort study.

### 1.2 Patients

This cohort was established on Feb 9, 2020 and the suspected and confirmed cases of COVID-19 admitted to the Xiangyang No.1 People’s Hospital Affiliated Hospital of Hubei University of Medicine according to the Diagnosis and Treatment Protocol for Novel Coronavirus Pneumonia (1st-7th editions) were included in this study ^[21]^. The retrospective data were traced back to Jan 22, 2020. The follow-up was carried out until Mar 28, 2020. Patients were classified into mild, moderate, severe, and critical type according to diagnosis and treatment protocol ^[21]^. The classification results were cross-checked by two experts. A third specialist involved if the inconsistency existed. This study was approved by the institutional review board (Approval number 2020GCP012). Informed consent from patients has been exempted since this study is an observational cohort study, which does not involve the personal privacy of patients nor incur greater than the minimal risk. Chinese clinical trial registry No.: ChiCTR2000031088.

### 1.3 Data collection

The clinical data of the confirmed COVID-19 cases were collected independently from the hospital information center and cross-checked, including the registry numbers, genders, ages, symptoms, time of symptom appearance and disappearance and outcomes. The day when the first symptom onset was designated as the day disease onset.

### 1.4 The method for symptoms quantized and the 43 symptoms

All symptoms over the whole course in the cohort were collected and the occurrence of a certain symptom was counted as one regardless of its frequency and intensity in that day while no symptom occurrence was counted as zero in that day, and the counted result was designated as the symptom frequency of that day for quantitative analysis in the further study. Every five-day from the onset day were designated as T1, T2, …, Tn, successively, where T1 was the first five-day period. In this cohort, the duration from disease onset to the day when 95% frequencies of all symptoms in the cohort disappeared was designated as the course and the duration from symptom onset to the day when all patients discharged or died was designated as the whole course.

The symptom occurrence rate was defined as the percentage of the cases who exhibited a certain symptom in the total cases in the cohort or in the different clinical types. The symptom proportion was defined as the percentage of the frequency of a certain symptom in the frequency of all symptoms in the cohort or in the different clinical types. The average frequency of all symptoms per case (AFAPC) of the cohort or the different clinical types over the course, the AFAPC within a certain day over the course and the average frequencies of all symptoms per case per day (AFAPCD) of the cohort or different clinical types over the course were calculated. The heat map of all symptoms in this cohort over the whole course since the symptom onset and the changes of frequency and AFAPC in all symptoms of the cohort or the clinical type was plotted.

The average frequency of a certain symptom per case (AFPC) and the average frequency of a certain symptom per case per day (AFPCD) over the course were calculated. And the number of cases exhibited a certain symptom, occurrence rate, proportion, AFPC, AFPCD, duration of symptom occurrence, time of frequency peak appearance, and frequency of peak value of each symptom were calculated. Temporal changes of the AFPC during periods before and after the moderate type and severe type transition to severe type and critical type were analyzed respectively. The changes during the periods from nine days before transition into severe type to three days after this transition in severe and critical cases, and from three days before transition into critical type to three days after this transition in critical cases were analyzed.

### 1.5 Classification, inter-correlation, difference, and association with clinical types in 22 symptoms

The 22 symptoms with occurring in number of patients ≥5 and frequency ≥20 were selected. These symptoms were identified for further analysis because the symptom with low occurrence rate or frequency was incapable for statistical analysis.

The 22 symptoms were classified and cross-checked independently by two experts according to the pathophysiological basis. Inconsistent results were solved by the third expert. For each patient, occurrence of a 22 symptom was scored 1 otherwise scored 0. A 0-1 matrix consisting of the number of 22 symptoms was set up and the inter-correlation coefficients of the 22 symptoms in the cohort was calculated. The results were illustrated by R corrplot. Coefficient ≥0.45 was statistically significant.

The occurrence rates of 22 symptoms between adjacent five-day periods over the course were analyzed, and statistically compared through Chi square test. *P*<0.05 was considered statistically significant.

The associations of the 22 symptoms with the moderate, severe, and critical types were analyzed through LASSO binomial logistic regression analysis. To discriminate between moderate type and the combined severe-critical type and between severe type and the critical type, SMOTE (Synthetic Minority Oversampling Technique) were applied through R SmoteClassif function with the Oversampling method for the combined group and the Undersampling method for the group of the moderate type. The multinomial logistic regression model was used to output the cases of the moderate, severe, and critical type. The output results accuracy was obtained by comparing the output results with the classification depended on the diagnostic and therapeutic guidelines of COVID-19 ^[21]^.

### 1.6 The 17 major symptoms, their temporal changes, and changing characteristics in the moderate, severe, and critical type

The 17 major symptoms obtained from the analysis including inter-correlation analysis, differences of occurrence rates between adjacent five-day periods and between moderate, severe, and critical type, association of symptoms with different clinical types and excluding the symptoms occurring in number of patients <10 and frequency <30. Among the major symptoms, we illustrated the changes of their frequencies per day in cohort, and the occurrence rates per five days in different clinical types.

### 1.7 Outcome

The outcome of cases were recorded and the changes of frequency in different outcomes were illustrated. Moreover, the correlation between outcome and major symptoms were analyzed through spearman correlation test.

### 1.8 Statistical analysis

All data were analyzed through SPSS20. Binary/dichotomous data were described by frequency or percentage of cases. The mean and standard deviation were calculated for the continuous data passing the normal distribution test. The median and the interquartile range (IQR) were used for those data sets that failed to pass the normal distribution test. Chi square test was applied to compare statistical significance of differences between groups. LASSO binomial logistic regression was used in a multinomial logistic regression model to identify association of symptoms with the cases of the moderate, severe, and critical types. The association between outcome and major symptoms was conducted through spearman correlation test.

## 2. Results

### 2.1 General information

A total of 542 suspected and confirmed cases, including 400 cases with negative results in nucleic acid tests and 142 cases with positive results, were admitted to our hospital before Feb 28, 2020. Among 142 cases, the data of 9 cases could not be traced back because they transferred from other hospitals, thus a total of 133 patients were included in this study. The basic information including: gender, age and the durations from symptom onset to hospital admission of the mild, moderate, severe, and critical types were listed in Table 1. There were 4, 89, 21, and 19 cases in the mild, moderate, severe, and critical type, respectively (Table 1). And difference in the ratio of male and female between moderate type and critical type was significant (*P*<0.05). Moreover, there were significant differences in age between moderate and severe type (*P*<0.001), moderate and critical type (*P*<0.001), and severe and critical type (*P*<0.001).

**Table 1.**
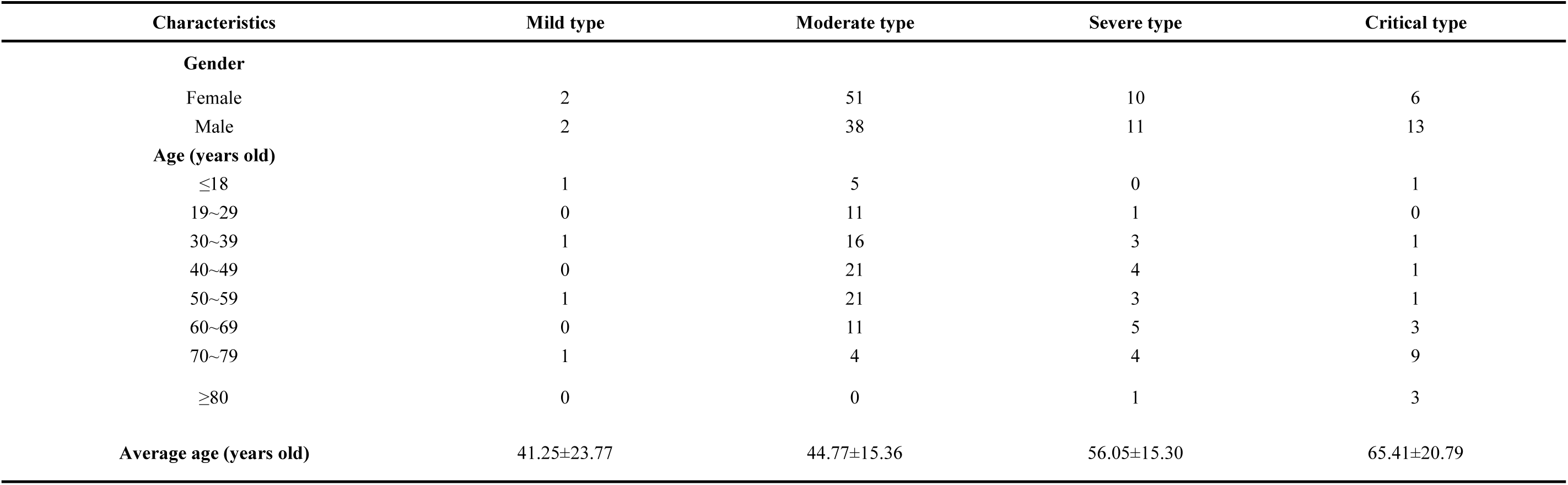
Baseline information of the cohort in the mild, moderate, severe, and critical types

### 2.2 All 43 symptoms and their daily changes in the cohort and the different clinical types, and changes during the transition of clinical type

#### 2.2.1 The symptoms and their daily change over the whole course in the cohort

According to our quantitative symptoms, a total of 43 symptoms and 6067 times of frequencies were identified in 133 patients during the whole course. The number of patients, and the occurrence rate, proportion, AFPC, AFPCD, the duration of symptom occurrence, the time of peak frequency appearance, the frequency peak value of symptoms were listed in Table 2. The 43 symptoms and the daily frequencies of each symptom in the whole course of COVID-19 were illustrated in Figure. 1. The results showed that the symptoms with occurrence rates>30.0% included fever (88.7%), poor appetite (81.2%), cough (81.2%), expectoration (54.9%), shortness of breath (44.4%), chest distress (42.1%), fatigue (39.8%), and diarrhea (35.3%) (Table 2). The symptoms with proportion>5.0% included cough (21.3%), fever (13.3%), poor appetite (13.1%), expectoration (8.8%), shortness of breath (5.9%), and chest distress (5.2%) (Table 2).

**Table 2.** All symptoms of the cohort and their occurrence rate, proportion, duration of symptom occurrence, and time of frequency peak appearance and the corresponding frequency peak value

| Symptom | Occurrence rate<br>(case) | Proportion<br>(Frequency) | AFPC | AFPCD | Duration of<br>symptom<br>occurrence (day) | Time of frequency<br>peak appearance (day) | Frequency<br>of peak<br>value |
| --- | --- | --- | --- | --- | --- | --- | --- |
| Fever | 88.7% (118) | 13.3%(808) | 6.85 | 0.43 | 1-16 | 1 | 96 |
| Poor appetite | 81.2% (108) | 13.1%(792) | 7.33 | 0.43 | 1-17 | 1 | 92 |
| Cough | 81.2% (108) | 21.3%(1293) | 11.97 | 0.39 | 1-31 | 1 | 76 |
| Expectoration | 54.9% (73) | 8.8%(535) | 7.33 | 0.22 | 1-33 | 3 | 33 |
| Shortness of breath | 44.4% (59) | 5.9%(360) | 6.10 | 0.18 | 1-34 | 1 | 22 |
| Chest distress | 42.1% (56) | 5.2%(317) | 5.66 | 0.20 | 1-28 | 2,3 | 25 |
| Fatigue | 39.8% (53) | 3.5%(215) | 4.06 | 0.19 | 1-21 | 3,8 | 16 |
| Diarrhea | 35.3% (47) | 3.4%(207) | 4.40 | 0.18 | 1-25 | 6,10,11 | 17 |
| Dyspnea | 27.8% (37) | 6.8%(415) | 11.22 | 0.30 | 1-37 | 13 | 18 |
| Nausea | 25.6% (34) | 2.8%(172) | 5.06 | 0.28 | 1-18 | 10 | 16 |
| Chills | 22.6% (30) | 2.7%(165) | 5.50 | 0.39 | 1-14 | 1 | 24 |
| Abdominal discomfort | 22.6% (30) | 2.3%(137) | 4.57 | 0.13 | 1-34 | 6 | 8 |
| Vomiting | 15.0% (20) | 1.5%(92) | 4.60 | 0.18 | 1-25 | 12 | 7 |
| pharyngeal discomfort | 12.8% (17) | 1.5%(94) | 5.53 | 0.55 | 1-10 | 1 | 13 |
| Palpitation | 12.0% (16) | 0.9%(57) | 3.56 | 0.14 | 1-26 | 9,10 | 4 |
| Dizziness | 9.8% (13) | 0.8%(48) | 3.69 | 0.15 | 1-24 | 6 | 5 |
| Acid reflux | 9.0% (12) | 0.5%(29) | 2.42 | 0.13 | 3-18 | 9,11,12 | 3 |
| Myalgia | 8.3% (11) | 0.8%(47) | 4.27 | 0.43 | 1-10 | 1 | 9 |
| Runny nose | 7.5% (10) | 1.0%(58) | 5.80 | 0.15 | 1-38 | 1,2,3 | 8 |
| Chest pain | 5.3% (7) | 0.5%(33) | 4.71 | 0.39 | 1-12 | 1,3,4,5,6 | 4 |
| Nasal obstruction | 5.3% (7) | 0.6%(36) | 5.14 | 0.57 | 1-9 | 1 | 6 |
| Sweat | 3.8% (5) | 0.2%(11) | 2.20 | 0.14 | 1-16 | 15 | 2 |
| Waist discomfort | 3.8% (5) | 0.4%(26) | 5.20 | 0.29 | 1-18 | 13 | 3 |
| Confusion | 3.0% (4) | 0.1%(7) | 1.75 | 0.11 | 11-16 | 13 | 2 |
| Headache | 3.0% (4) | 0.4%(23) | 5.75 | 0.82 | 1-7 | 1 | 4 |
| Dry mouth | 3.0% (4) | 0.2%(13) | 3.25 | 0.30 | 1-11 | 7,10,11 | 2 |
| Heartburn | 2.3% (3) | 0.1%(7) | 2.33 | 0.19 | 6-12 | 10,11 | 2 |
| Bleeding gums | 1.5% (2) | 0.05%(3) | 1.50 | 0.30 | 1-5 |  | 1 |
| Hematuresis | 1.5% (2) | 0.1%(6) | 3.00 | 0.23 | 1-13 |  | 1 |
| Bitter taste | 1.5% (2) | 0.1%(8) | 4.00 | 0.29 | 2-14 |  | 1 |
| Shoulder pain | 1.5% (2) | 0.3%(20) | 10.00 | 0.56 | 1-18 | 10,12 | 2 |
| Nasal bleeding | 1.5% (2) | 0.08%(5) | 2.50 | 0.14 | 6-18 |  | 1 |
| Colporrhagia | 0.8% (1) | 0.03%(2) | 2.00 |  |  |  | 1 |
| Ocular discomfort | 0.8% (1) | 0.08%(5) | 5.00 |  |  |  | 1 |
| Chest discomfort | 0.8% (1) | 0.03%(2) | 2.00 |  |  |  | 1 |
| Vulvar discomfort | 0.8% (1) | 0.02%(1) | 1.00 |  |  |  | 1 |
| Hand tremble | 0.8% (1) | 0.02%(1) | 1.00 |  |  |  | 1 |
| Depressed | 0.8% (1) | 0.03%(2) | 2.00 |  |  |  | 1 |
| Skin itch | 0.8% (1) | 0.03%(2) | 2.00 |  |  |  | 1 |
| Sneeze | 0.8% (1) | 0.05%(3) | 3.00 |  |  |  | 1 |
| Odynuria | 0.8% (1) | 0.1%(7) | 7.00 |  |  |  | 1 |
| Anabole | 0.8% (1) | 0.02%(1) | 1.00 |  |  |  | 1 |
| Thigh discomfort | 0.8% (1) | 0.03%(2) | 2.00 |  |  |  | 1 |
| Total symptoms |  | 100.0%(6067) | 45.62 | 1.57 | 1-29 | 1 | 463 |

AFPC: Average frequency of a certain symptom per case
AFPCD: Average frequency of a certain symptom per case per day

**Figure 1.**
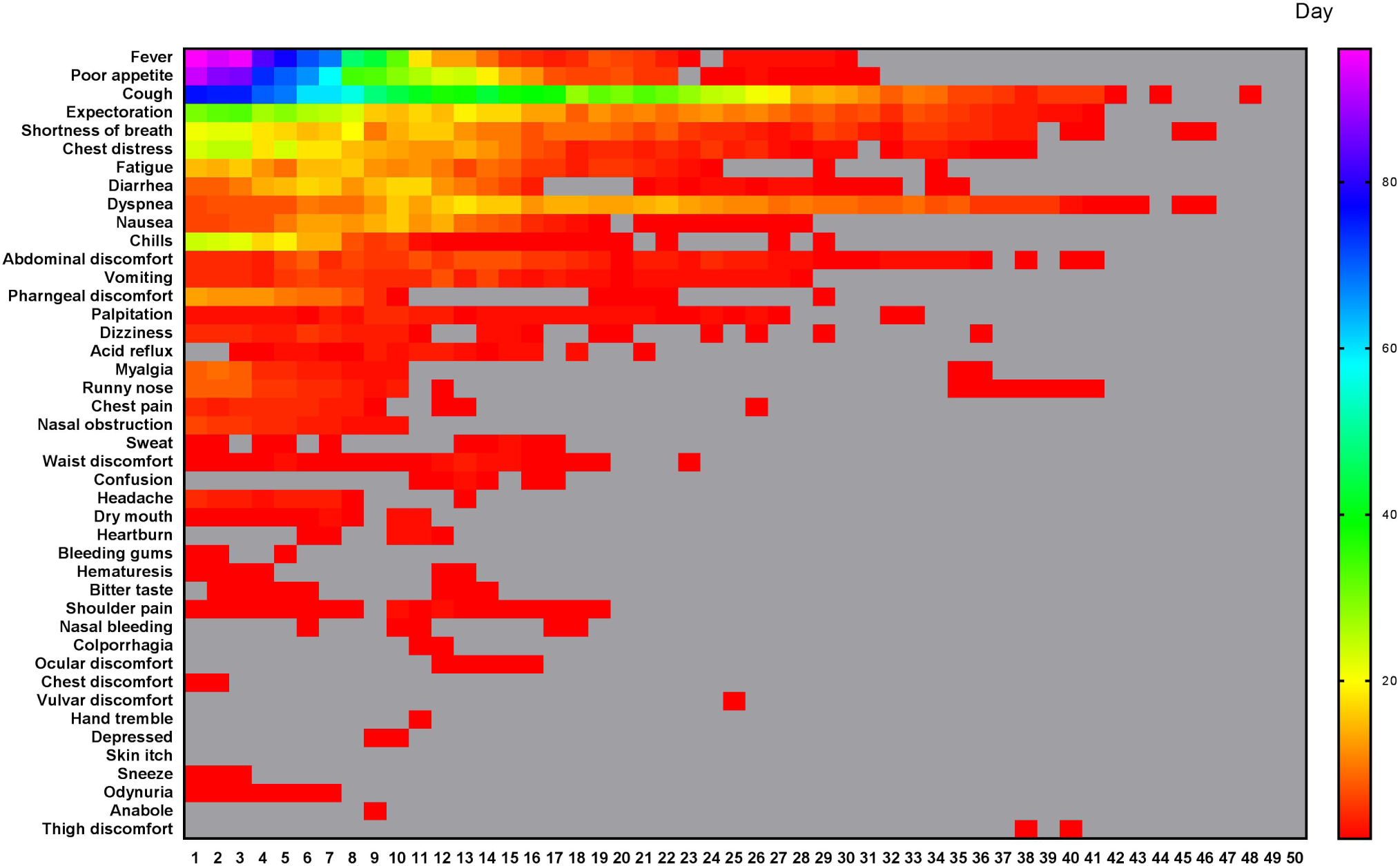
All different symptoms and their changes of frequency per day in the whole course of this cohort. A total of 43 symptoms were listed. The horizontal axis is the course time day. Color scale illustrates the magnitude of frequency. Gray block: the frequency is 0.

The daily frequency (Figure. 2A) and AFAPC (Figure. 2F) of all symptoms were plotted, which showed the changes of the 43 symptoms during the whole course since disease onset. The 95% frequencies of all symptoms disappeared on the day 29 since disease onset.

**Figure 2.**
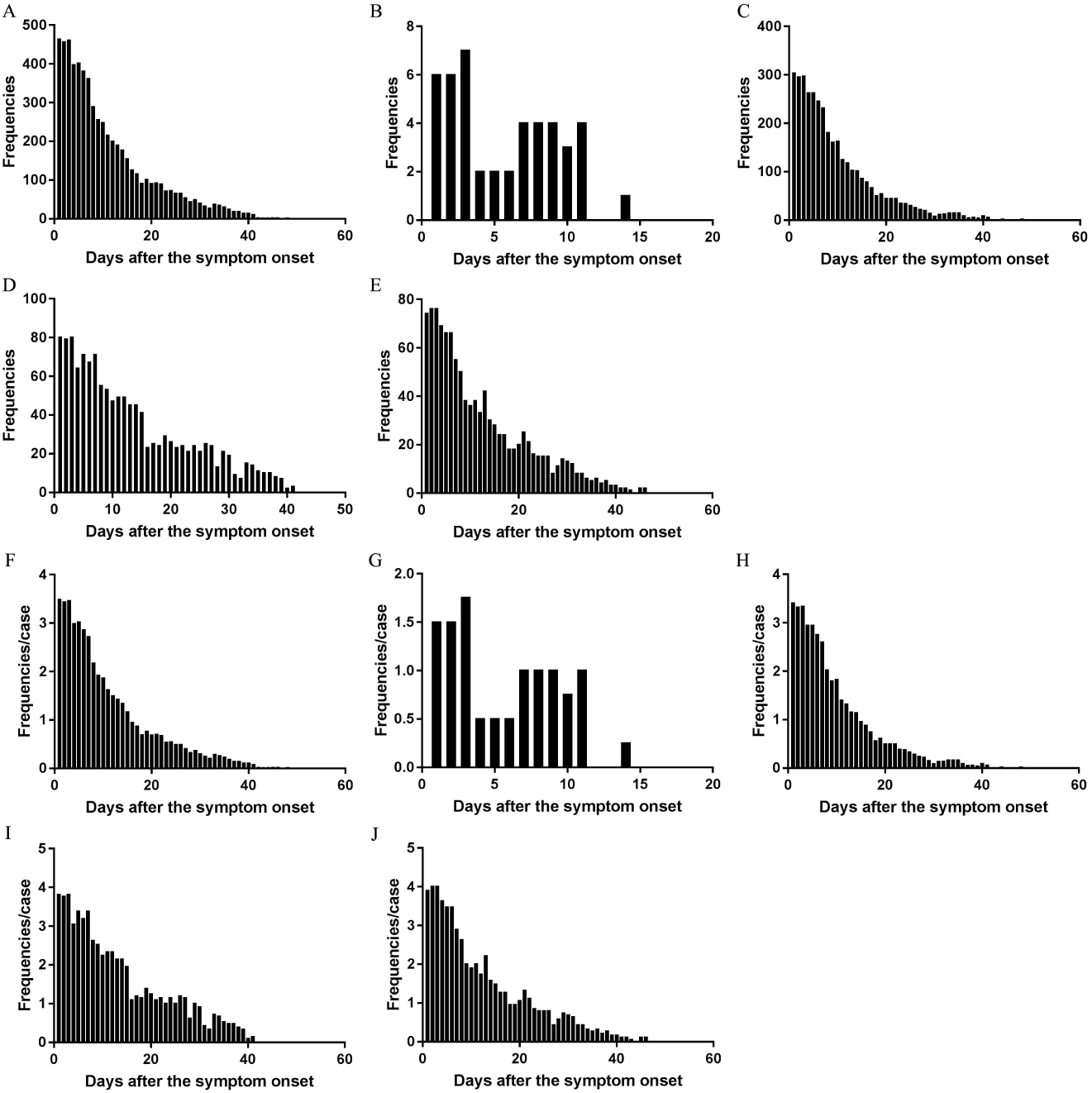
The changes of total frequencies and AFAPC of all symptoms in the cohort and in the different clinical types. The changes of frequency of all symptoms in the cohort (A), the mild type (B), the moderate type (C), the severe type (D), and the critical type (E). The changes of AFPC in the cohort (F), the mild type (G), the moderate type (H), the severe type (I), and the critical type (J).

#### 2.2.2 The symptoms and their daily changes over the whole course in the different clinical types

The daily changes of the total symptom frequencies and AFAPC in mild, moderate, severe, critical type had similar profiles (Figure. 2).

##### Mild type

There were 5 symptoms which occurred 45 times, with AFAPC as 11.25 and AFAPCD as 1.02 (Table 3). The peak frequency was on the first 3 days, with an average 6 times of frequencies per day. The 95% of the symptoms disappeared on the day 11 (Figure. 2B and 2G). Symptoms with occurrence rate >30.0% including: cough (100.0%), expectoration (75.0%), and fever (50.0%); Symptoms with proportion >10.0% including: cough (60.0%), expectoration (15.6%), fever (13.3%) (Table 3).

**Table 3.** The AFAPC, AFAPCD and the symptoms with occurrence rate > 30.0% in the mild, moderate, severe, and critical types

| Item | Mild type |  | Moderate type |  | Severe type |  | Critical type |
| --- | --- | --- | --- | --- | --- | --- | --- |
| AFPC | 11.25 |  | 40.28 |  | 63.52 |  | 58.05 |
| AFPCD | 1.02 |  | 1.55 |  | 1.92 |  | 1.81 |

| Symptom | Occurrence rate (case) | Proportion (frequency) | Occurrence rate (case) | Proportion (frequency) | Occurrence rate (case) | Proportion (frequency) | Occurrence rate (case) | Proportion (frequency) |
| --- | --- | --- | --- | --- | --- | --- | --- | --- |
| Fever | 50.0% (2) | 13.3% (6) | 87.6% (78) | 14.4% (517) | 95.2% (20) | 11.7% (156) | 94.7% (18) | 11.7% (129) |
| Poor appetite | 25.0% (1) | 6.7% (3) | 77.5% (69) | 12.6% (452) | 95.2% (20) | 14.4% (192) | 94.7% (18) | 13.1% (145) |
| Cough | 100.0% (4) | 60.0% (27) | 76.4% (68) | 23.4% (838) | 95.2% (20) | 21.4% (285) | 84.2% (16) | 13.0% (143) |
| Expectoration | 75.0% (3) | 15.6% (7) | 46.1% (41) | 8.6% (310) | 66.7% (14) | 9.1% (122) | 78.9% (15) | 8.7% (96) |
| Fatigue |  |  | 38.2% (34) | 4.2% (149) | 57.1% (12) | 2.6% (35) | 36.8% (7) | 2.8% (31) |
| Diarrhea |  |  | 32.6% (29) | 3.7% (131) | 42.9% (9) | 2.8% (37) | 47.4% (9) | 3.5% (39) |
| Chest distress |  |  | 31.5% (28) | 4.6% (165) | 57.1% (12) | 6.4% (86) | 84.2% (16) | 6.0% (66) |
| Shortness of breath |  |  | 27.0% (24) | 3.7% (134) | 81.0% (17) | 10.0% (134) | 94.7% (18) | 8.3% (92) |
| Chills |  |  | 22.5% (20) | 3.1% (112) | 33.3% (7) | 2.7% (36) | 15.8% (3) | 1.5% (17) |
| Abdominal discomfort |  |  | 21.3% (19) | 3.0% (107) | 14.3% (3) | 0.9% (12) | 42.1% (8) | 1.6% (18) |
| Dyspnea |  |  | 9.0% (8) | 1.3% (45) | 52.4% (11) | 8.1% (108) | 94.7% (18) | 23.8% (262) |

##### Moderate type

There were 39 symptoms which occurred 3585 times, with AFAPC as 40.28 and AFAPCD as 1.55 (Table 3). The peak frequency was on the first day with 303 times of frequencies. The 95% of the symptoms disappeared on the day 26 (Figure 2C and 2H). Symptoms with occurrence rate >30.0% including: fever (87.6%), poor appetite (77.5%), cough (76.4%), expectoration (46.1%), fatigue (38.2%), diarrhea (32.6%), and chest distress (31.5%); Symptoms with proportion >10.0% including: cough (23.4%), fever (14.4%), and poor appetite (12.6%) (Table 3).

##### Severe type

There were 27 symptoms which occurred 1334 times, with AFAPC as 63.52 and AFAPCD as 1.92 (Table 3). The peak frequency was on the first 3 days, with an average 79 times of frequencies per day. The 95% of the symptoms disappeared on the day 33 (Figure 2D and 2I). Symptoms with occurrence rate >30.0% including: fever (95.2%), poor appetite (95.2%), cough (95.2%), shortness of breath (81.0%), expectoration (66.7%), fatigue (57.1%), chest distress (57.1%), dyspnea (52.4%), diarrhea (42.9%), and chills (33.3%); Symptoms with proportion >10.0% including : cough (21.4%), poor appetite (14.4%), fever (11.7%), and shortness of breath (10.0%) (Table 3).

##### Critical type

There were 21 symptoms which occurred 1103 times, with AFAPC as 58.05 and AFAPCD as 1.81 (Table 3). The peak frequency was on the first 3 days, with an average 75 times of frequencies per day. The 95% of the symptoms disappeared on the day 32 (Figure 2E and 2J). Symptoms with occurrence rate >30.0% including: fever (94.7%), poor appetite (94.7%), shortness of breath (94.7%), dyspnea (94.7%), cough (84.2%), chest distress (84.2%), expectoration (78.9%), diarrhea (47.4%), abdominal discomfort (42.1%), and fatigue (36.8%); Symptoms with proportion >10.0% including : dyspnea (23.8%), poor appetite (13.1%), cough (13.0%), and fever (11.7%) (Table 3).

#### 2.2.3 Changes of symptoms before and after transition into the severe and critical types

The duration from symptom onset to transition into the severe type in severe and critical cases was 8.15 ± 5.29 days. The durations from symptom onset to transition into the severe type in severe cases was 8.67±5.22 days, and in critical cases was 7.56 ± 5.30 days, but there was no significant difference between the two types (Figure 3A). The duration from the severe to transition into the critical type in critical cases was 4.11 ± 5.65 days.

**Figure 3.**
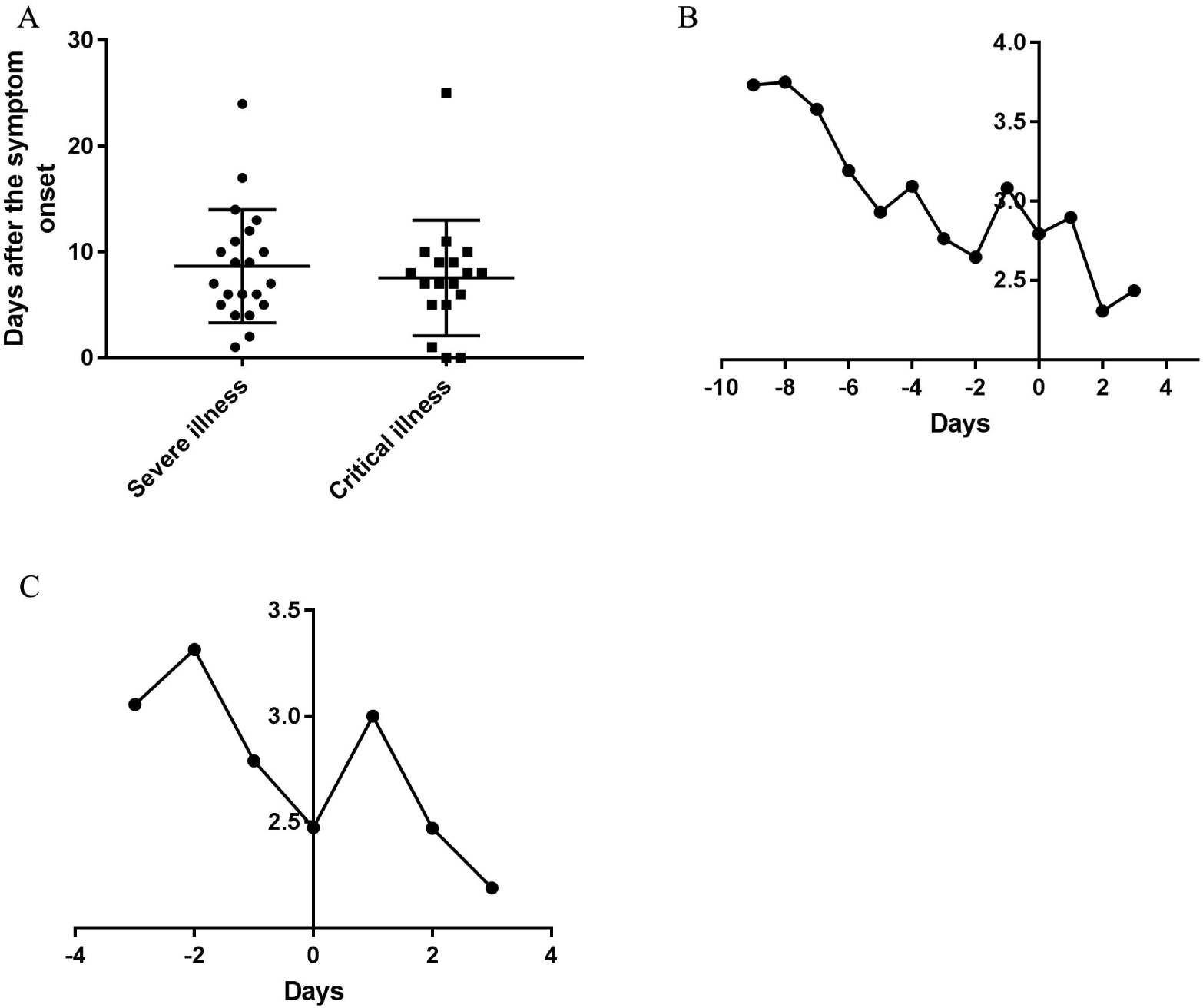
Durations of the courses from onset to transition into the severe and critical types and temporal changes of AFPC of symptoms before and after transition. (A) Durations of the courses from onset to transition into the severe and critical types. (B) The changes of AFPC in the severe and critical cases before and after transition into severe type. (C) The changes of AFPC in the critical cases before and after transition into critical type. 0 in the horizontal axis represents the transition date of the severe type developed from moderate type (B) and the transition date of the critical type developed from severe type (C), respectively.

The daily AFAPC in severe and critical cases was decreased continuously 9 days before the transition into the severe type and suddenly increased on the day 2 before the day transition into the severe type (Figure 3B); Correspondingly, it decreased continuously before the day transition into the critical type and then increased from the transition day of the critical type to the next day and then decreased (Figure 3C).

### 2.3 Analysis of 22 symptoms in difference, inter-correlation, association, and classification these symptoms based on pathophysiology

For the symptom with low occurrence rate or frequency was incapable for statistical analysis, we selected 22 symptoms with each occurring in number of patients ≥5 and frequency ≥20 among the 43 symptoms in the following study to analyze the symptoms systematically.

#### 2.3.1 Pathophysiology-based classification and inter-correlation analysis of symptoms

The results of pathophysiology-based classification showed that 22 symptoms were classified into three categories: (i) Upper respiratory tract infection symptoms: fever, chills, nasal obstruction, runny nose, myalgia, dizziness, waist discomfort, pharyngeal discomfort, and fatigue; (ii) cardiopulmonary symptoms: cough, expectoration, shortness of breath, dyspnea, and chest distress, palpitation, and chest pain; (iii) gastrointestinal symptoms: poor appetite, nausea, vomiting, diarrhea, abdominal discomfort, and acid reflux.

The results of inter-correlation analysis showed high correlation between cough and expectoration, shortness of breath and dyspnea, nausea and vomiting, shortness of breath and chest distress, dyspnea and chest distress, nasal obstruction and runny nose, nasal obstruction and myalgia, and between runny nose and myalgia (Figure 4, Supplemental Table 1).

**Figure 4.**
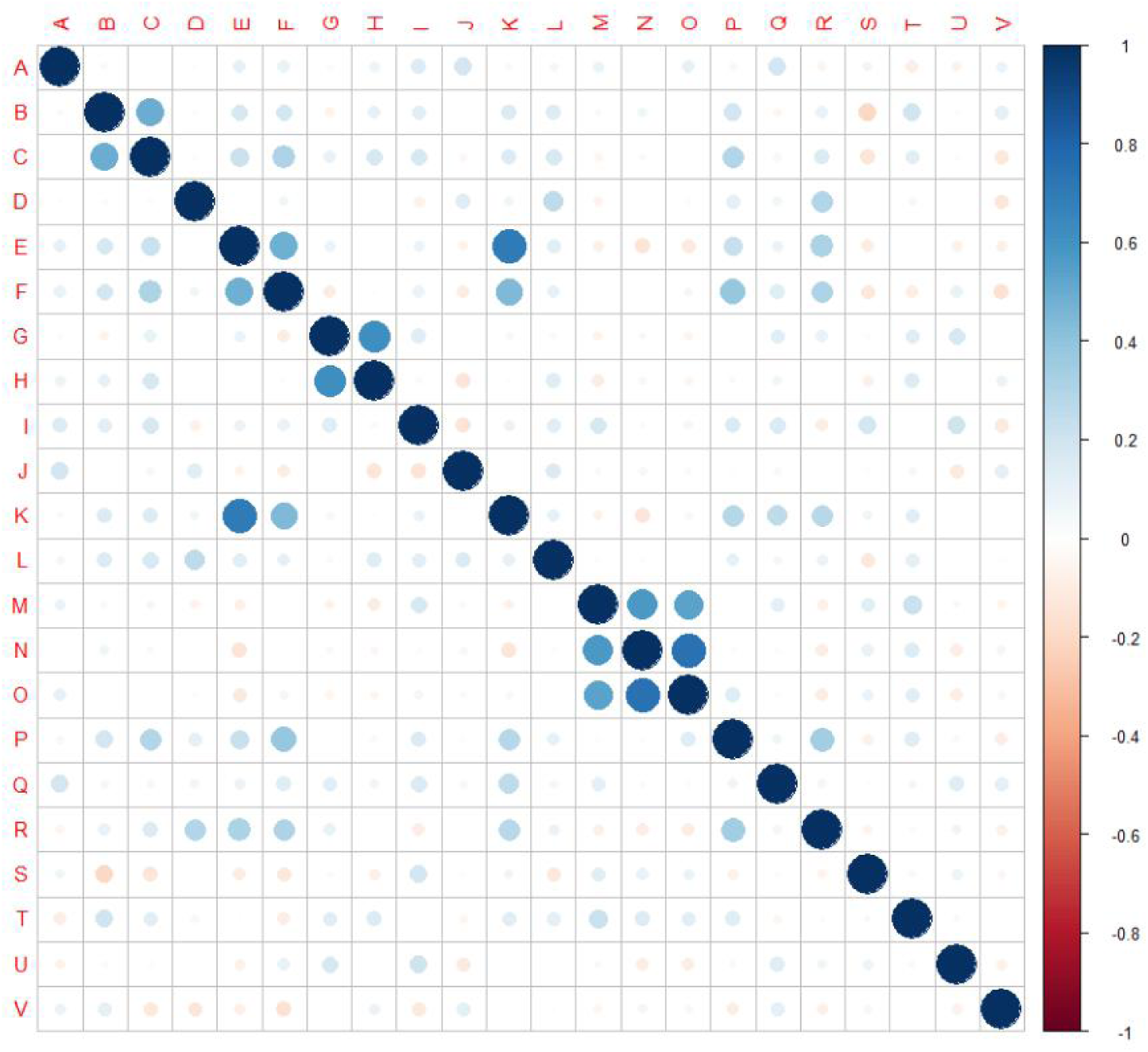
Inter-correlation among 22 symptoms. The 22 symptoms in the cohort were identified and analyzed their inter-correlation. (A) fever, (B) cough, (C) expectoration, (D) fatigue, (E) shortness of breath, (F) dyspnea, (G) nausea, (H) vomiting, (I) diarrhea, (J) chills, (K) chest distress, (L) poor appetite, (M) nasal obstruction, (N) runny nose, (O) myalgia, (P) palpitation, (Q) abdominal discomfort, (R) dizziness, (S) waist discomfort, (T) pharyngeal discomfort, (U) Acid reflux, (V) chest pain. Scale bar: Positive value, positive correlation. Negative value, negative correlation. Larger absolute value of the value represents higher correlation.

#### 2.3.2 Differences in the occurrence rates of symptoms between clinical types and between adjacent five-day periods over the course

We compared the occurrence rate of 22 symptoms among the moderate type, severe type, and critical type in 129 cases. The occurrence rates of fatigue (*P*<0.001), shortness of breath (*P*<0.001), dyspnea (*P*<0.001), chest distress (*P*<0.05), palpitation (*P*<0.01), and dizziness (*P*<0.01) were higher in severe type than those in moderate type. The occurrence rates of expectoration (*P*<0.01), shortness of breath (*P*<0.001), dyspnea (*P*<0.001), chest distress (*P*<0.001), palpitation (*P*<0.01), and dizziness (*P*<0.05) were higher in critical type than those in moderate type. The occurrence rates of dyspnea (*P*<0.001) and abdominal discomfort (*P*<0.05) were higher, and fatigue (*P*<0.05) was lower in critical type than those in severe type. Overall, a total of 8 symptoms (expectoration, fatigue, shortness of breath, dyspnea, chest distress, palpitation, abdominal discomfort, and dizziness) exhibited significant difference (*P*<0.05) in the occurrence rates among three clinical types.

We analyzed differences in the occurrence rates of 22 symptoms between adjacent periods over the course. The occurrence rates of fever (*P*<0.01), poor appetite (*P*<0.01), and myalgia (*P*<0.05) were higher, and dyspnea (*P*<0.01), nausea (*P*<0.05), and diarrhea (*P*< 0.05) were lower in T1 than those in T2; The occurrence rates of fever (*P*<0.001), chills (*P*<0.001), poor appetite (*P*< 0.001), and pharyngeal discomfort (*P*<0.01) were higher in T2 than those in T3; The occurrence rates of fever (*P*<0.05), fatigue (*P*<0.05), nausea (*P*<0.05), diarrhea (*P*<0.001), and poor appetite (*P*< 0.05) were higher in T3 than those in T4; The occurrence rates of cough (*P* <0.05) was higher in T6 than T7; The occurrence rates of expectoration (*P*<0.05) was higher in T8 than those in T9. Overall, a total of 11 symptoms (fever, cough, expectoration, fatigue, dyspnea, nausea, diarrhea, chills, poor appetite, myalgia, and pharyngeal discomfort) were significantly different between periods over the course.

#### 2.3.3 Analysis of the association in symptoms with the clinical types based on the LASSO binomial logistic regression

Among the 22 symptoms, expectoration, shortness of breath, dyspnea, diarrhea, and poor appetite were positively correlated with the combined severe-critical type, and vomiting, waist discomfort, pharyngeal discomfort and acid reflux were negatively correlated with the combined severe-critical type, when discriminating between the moderate type and the combined type (Supplemental Table 2). The accuracy output of multinomial logistic regression model through these indentified symptoms discriminate the moderate type and the combined severe-critical type was 84.5%, and discriminate the severe type and critical type was 70.0%, only dyspnea was positively correlated with the critical type (Supplemental Table 3). A total of 9 symptoms (expectoration, shortness of breath, dyspnea, diarrhea, poor appetite, vomiting, waist discomfort, pharyngeal discomfort, acid reflux) were significantly associated with the clinical type.

### 2.4 The screened 17 major symptoms, their temporal changes, and characteristics in the different clinical types

#### 2.4.1 Major symptoms

According to the analysis of 22 symptoms including inter-correlation analysis, differences of occurrence rates between adjacent five-day periods and between moderate, severe, and critical type, and association of symptoms with different clinical types, we identified 20 symptoms firstly. And then we excluding the symptoms occurring in number of patients ≤10 and frequencies ≤ 30, namely excluding the nasal obstruction, runny nose and waist discomfort. A total of 17 major symptoms including fever, poor appetite, cough, expectoration, shortness of breath, chest distress, fatigue, diarrhea, dyspnea, nausea, chills, abdominal discomfort, vomiting, pharyngeal discomfort, palpitation, dizziness, and myalgia were obtained finally.

#### 2.4.2 Temporal changes of the major symptoms and their changes in the different clinical types

The changes of frequencies (Figure 5), and occurrence rates in moderate, severe, and critical type (Figure 6) were illustrated. The results showed that the trend were similar in the temporal changes of occurrence rate in fever, poor appetite, fatigue, diarrhea, nausea, chills, pharyngeal discomfort, and myalgia among the clinical types (Figure 6). Fever occurred in 1-16 days and poor appetite occurred in 1-17 days. Their frequencies were the highest on the onset day, and gradually decreasing thereafter (Figure 5). Fatigue occurred in 1-21 days and the peak value of its frequency was distributed on the day 3 and day 8. Diarrhea occurred in 1-25 days and the peak value was on the day 6, day 10, and day 11. Nausea occurred in 1-18 days and the peak value was on the day 10. Chills occurred in 1-14 days, pharyngeal discomfort in 1-10 days and myalgia occurred in 1-10 days. Their frequencies were the highest on the onset day, and decreased dramatically afterwards (Figure 5).

**Figure 5.**
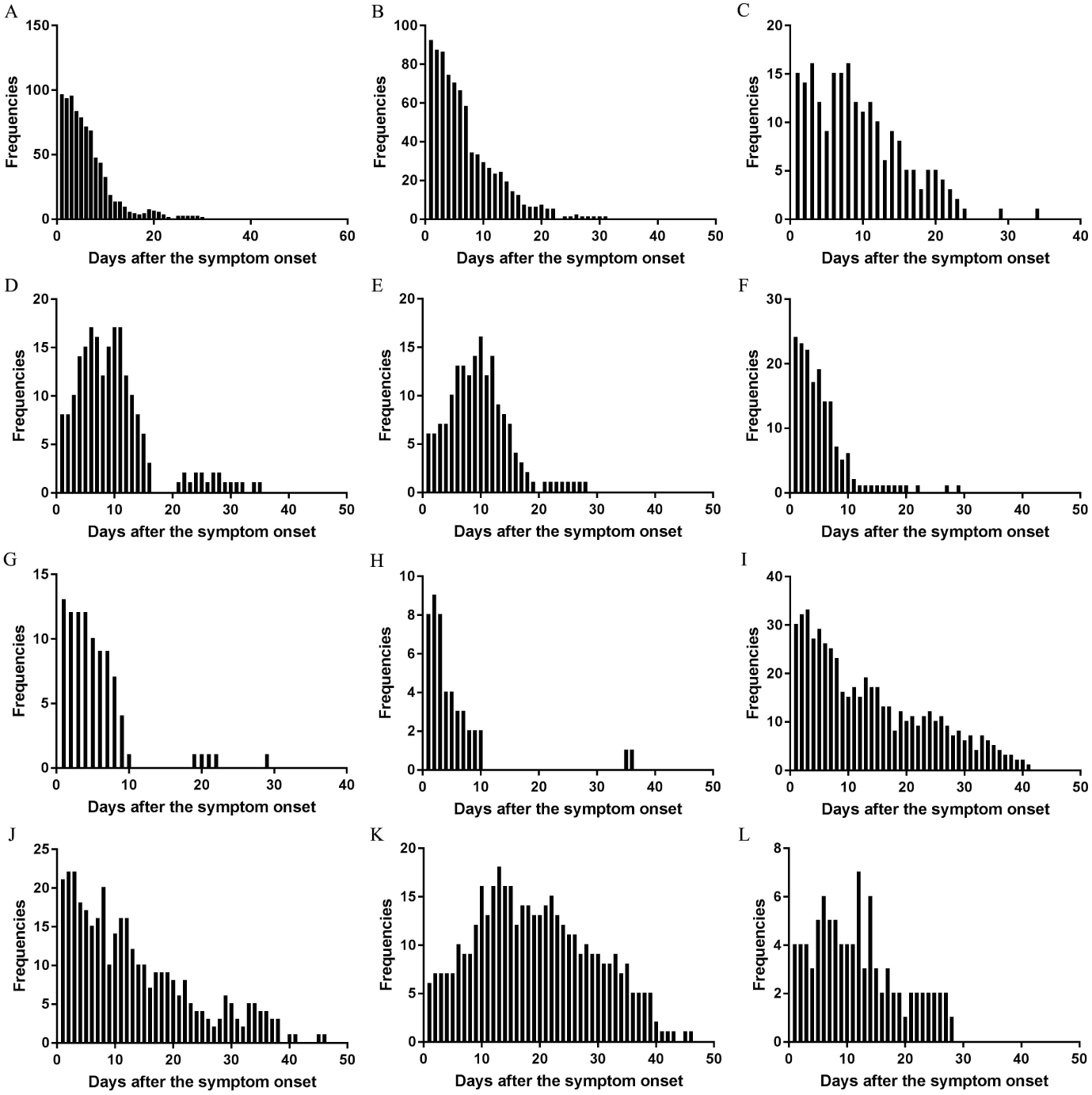
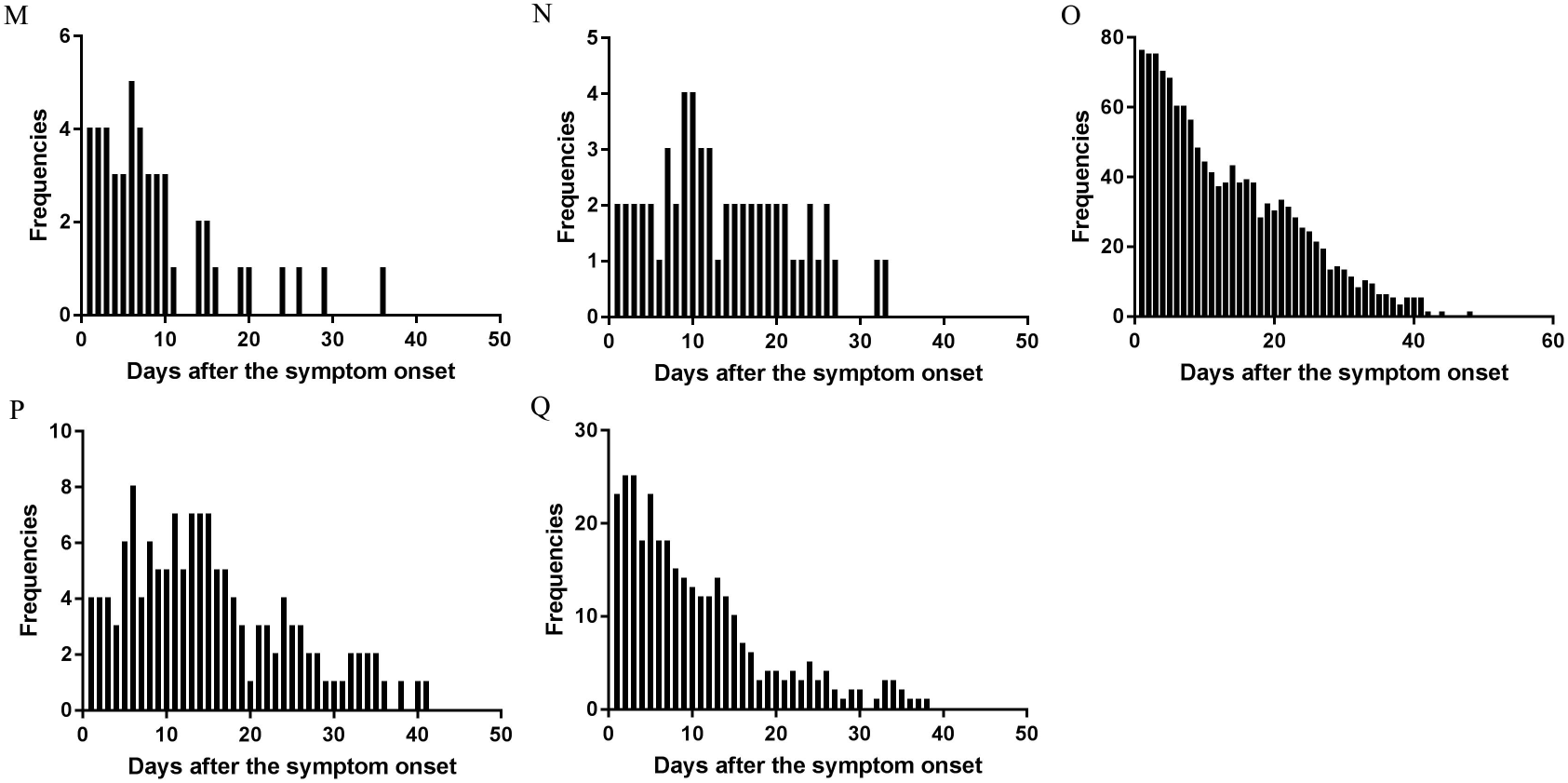
The changes of frequency of major symptoms over the whole course in the cohort. (A) Fever, (B) Poor appetite, (C) Fatigue, (D) Diarrhea, (E) Nausea, (F) Chills, (G) Pharyngeal discomfort, (H) Myalgia, (I) Expectoration, (J) Shortness of breath, (K) Dyspnea, (L) Vomiting, (M) Dizziness, (N) Palpitation, (O) Cough, (P) Abdominal discomfort, (Q) Chest distress.

**Figure 6.**
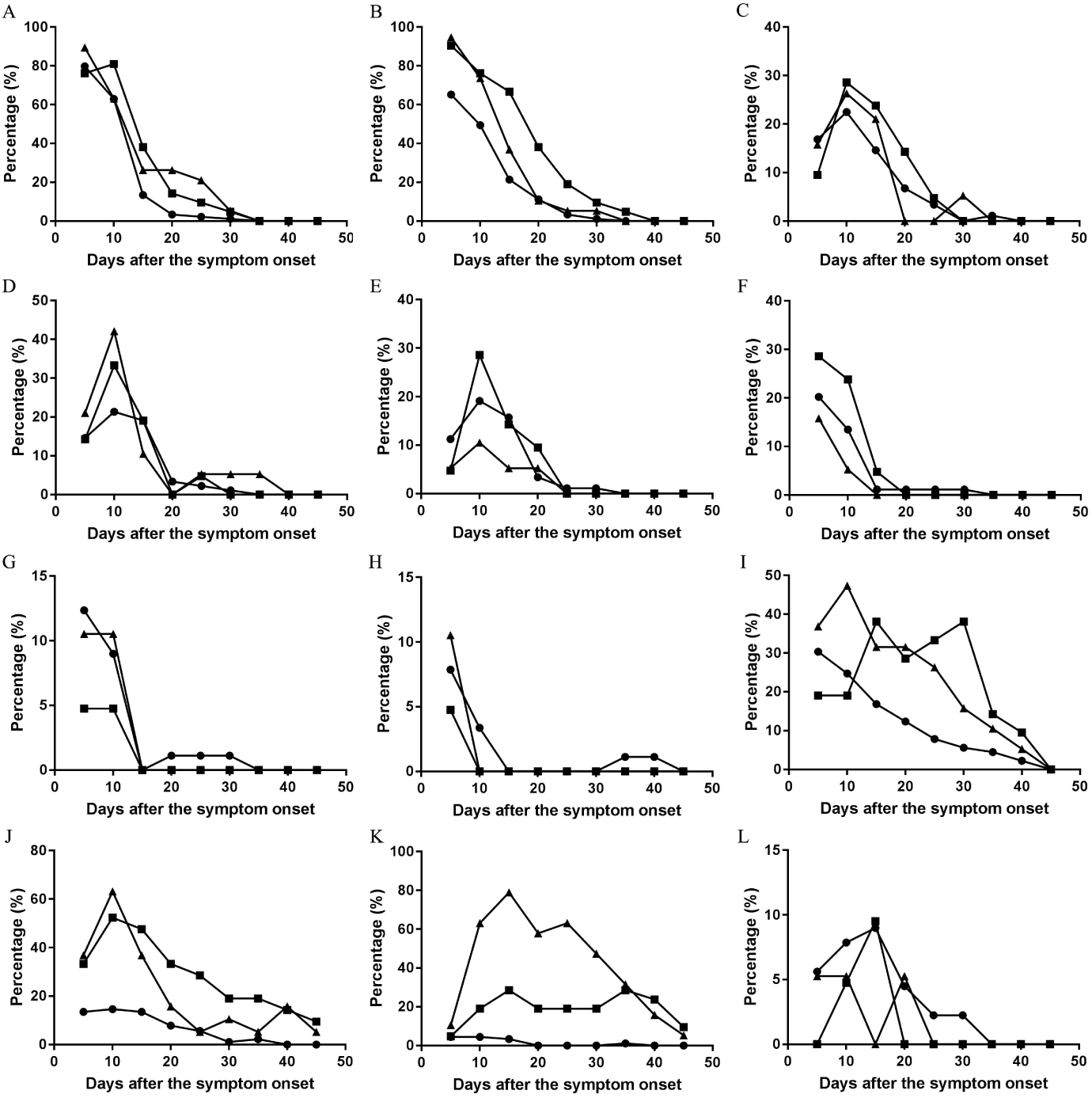
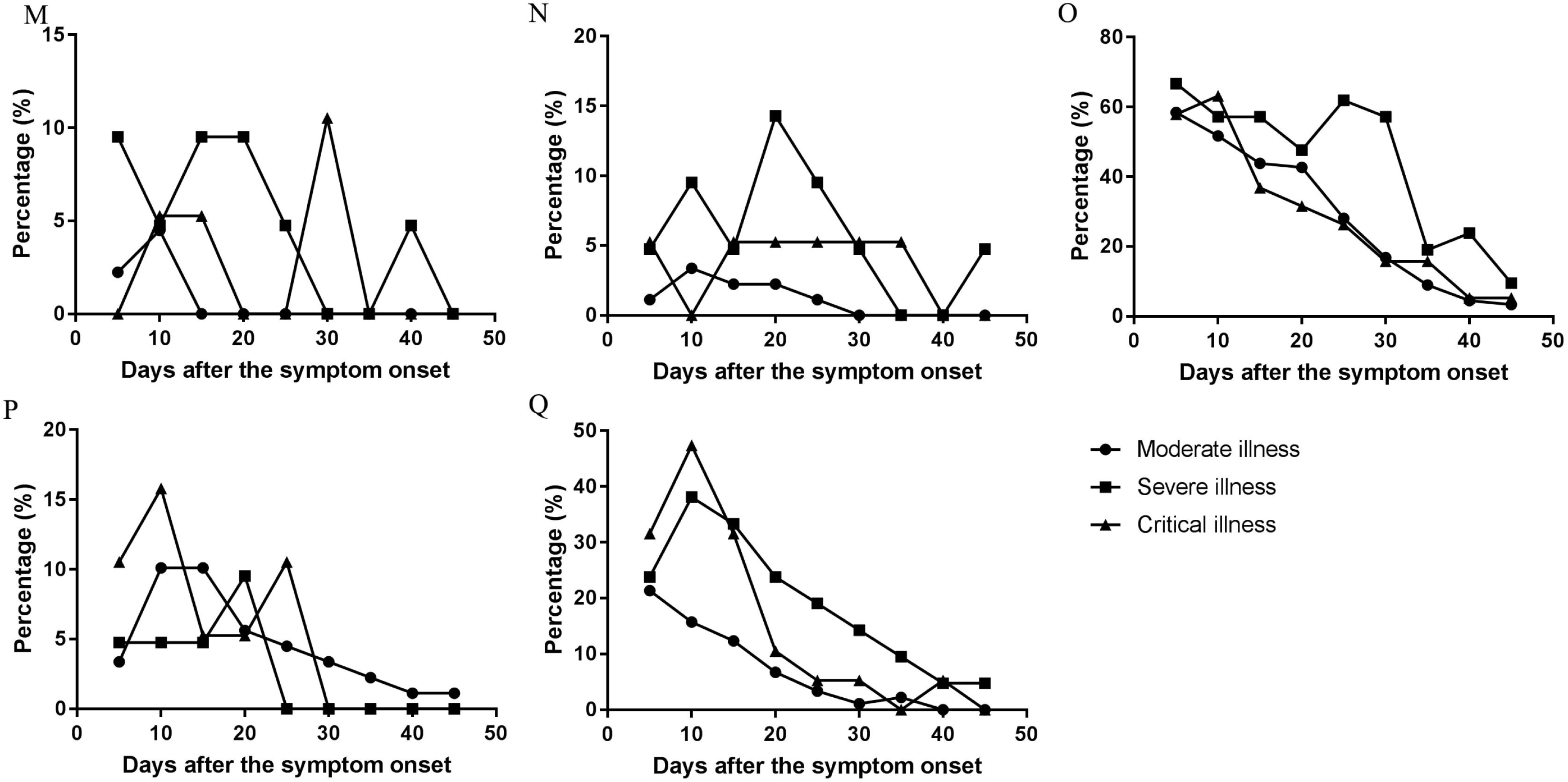
The changes of occurrence rate of major symptoms per five day over the whole course in the moderate, severe, and critical types. (A) Fever, (B) Poor appetite, (C) Fatigue, (D) Diarrhea, (E) Nausea, (F) Chills, (G) Pharyngeal discomfort, (H) Myalgia, (I) Expectoration, (J) Shortness of breath, (K) Dyspnea, (L) Vomiting, (M) Dizziness, (N) Palpitation, (O) Cough, (P) Abdominal discomfort, (Q) Chest distress.

There were differences in the temporal change trend of the occurrence rates of expectoration, shortness of breath, dyspnea, vomiting, dizziness, palpitation, cough, abdominal discomfort, chest distress among the moderate, severe, and critical types (Figure 6). Expectoration occurred in 1-33 days and its frequency was the highest three days since symptom onset (Figure 5). Shortness of breath occurred in 1-34 days and its frequency was the highest on the onset day. Dyspnea occurred in 1-37 days and the peak value of its frequency was on the day 13. Vomiting occurred in 1-25 days and the peak value of its frequency was on the day 12. Dizziness occurred in 1-24 days and the peak value of its frequency was on the day 6. Palpitation occurred in 1-26 days and the peak value of its frequency was on the day 9 and day 10. Cough occurred in 1-31 days and its frequency was the highest on the onset day (Figure 5). The temporal change of the occurrence rate of cough in the severe type was different from those in moderate or critical type (Figure 6). Abdominal discomfort occurred in 1-34 days and the peak value of its frequency was on the day 6. The temporal change of the occurrence rate of abdominal discomfort in the critical type was different from the moderate or severe type (Figure 6). Chest distress occurred in 1-28 days and its frequency was the highest in the first three days after onset. The temporal change of the occurrence rate of chest distress in the moderate type was different from the severe or critical type (Figure 6).

### 2.5 Outcomes

As of Mar 28, 2020, there were 123 patients discharged from the hospital and 10 died, the mortality rate was 7.52%. The average age of the death was 73.10±8.99 years old. The correlation was found between death and dyspnea (*P*<0.001), shortness of breath (*P*<0.01) and chest distress (P<0.05) among 17 major symptoms. The coefficient of them was 0.393, 0.258, 0.214, respectively.

## Discussion

The comparisons in this cohort indicated that the age might associated with the severity of the illness and the number of the male in critical type was higher than those in moderate type. But for the reason that the mild cases just contain 4 cases, thus we couldn’t conduct statistical analysis on them. In this study, we transformed the descriptive data into binary categorical data, performed a quantitative analysis and showed the changes and characteristic over the whole course of all symptoms of COVID-19 cases in this cohort through heat map and detailed table. Furthermore, the results of inter-correlation analysis between symptoms, and output accuracy obtained from multinomial logistic regression model, supporting that our quantitative analysis of symptoms is credible.

Temporal changes of frequencies in total symptoms indicated that the frequencies were the highest one week after symptom onset, 74% symptoms frequencies disappeared in two weeks. On the day 29 after onset, 95% all symptoms frequencies disappeared and only the symptoms like cough, expectoration and respiratory failure were present. Consistent results were observed from the comparisons between symptoms in adjacent five-day periods. The significantly different changes mostly occurred in the first 20 days after disease onset. Therefore, the symptoms mainly occurred in the first 20 days since symptom onset, few symptoms occurred in 20-30 days, and only cough and expectoration existed 30 days after onset.

The symptoms, occurrence rates, proportion and disappearing time of symptoms differ in different clinical types. The number of patients and symptoms, and disease course of mild type were the fewest among clinical types. Our data showed that the AFPC and the AFPCD of the course in four clinical types exhibited that the severe type> critical type> moderate type> mild type. Similarly, the duration of disease course from onset to the time when 95% symptoms frequencies disappear also showed a rule that the severe type (33 d)> critical type (32 d)> the moderate type (26 d)> mild type (11 d). Studies have showed that hospitalization durations of the death cases which belong to the critical type are short (14.70 ± 9.60 days) ^[22, 23]^, partly accounting for shorter duration of disease course of the critical type than the severe type. In addition, the dominant symptom in the critical type is dyspnea ^[24]^, which necessitates respiration assisting equipment especially after the occurrence of consciousness disorders and may mask other symptoms. Therefore, more kinds of symptoms and the higher frequency of symptoms, and longer course of disease might be associated with more serious disease.

We have analyzed the association of symptoms with the clinical types through LASSO binomial logistic regression. These identified symptoms can be used to discriminate the patients in the different clinical types, among them only dyspnea was positively correlated with the critical type and can be used to discriminate patients between severe and critical type. Moreover, the study in the correlation between outcome and major symptoms suggested that dyspnea, shortness of breath, chest distress were correlated with death.

Comparison of the occurrence rates of symptoms per five days showed that there were differences in expectoration, shortness of breath, dyspnea, vomiting, and palpitation. The occurrence rates of dyspnea and palpitation were high and their durations were long in severe and critical cases. Palpitation mainly occurred in severe cases, and the recovery time was shorter than dyspnea, suggesting that palpitation may be more closely related to heart injury for the reason that palpitation usually associated with respiratory disorder or heart injury. The increase of abdominal discomfort around 20 days and 25 days was related to the increase of digestive complications. In the early stage of COVID-19, the main symptoms are fever, poor appetite, and cough in various clinical types. These main symptoms are associated with upper respiratory tract infection and similar to the general reaction and discomfort caused by common virus infection ^[25]^, but not specific to COVID-19. The occurrence rate of vomiting, dizziness and abdominal discomfort are different in the early stage of the disease. In later stages, the severe cases have other cardiopulmonary symptoms such as expectoration, shortness of breath, fatigue, dyspnea and chest distress, which indicates that the lung function is seriously damaged and enters the decompensated period. The main symptoms in the critical cases include dyspnea, chest distress, expectoration and diarrhea, where dyspnea is the dominant one with highest occurrence rate and highest proportion in the clinical type. Therefore, there is no characteristic symptom for each types in early stage. The severe type is characterized by multiple symptoms related to respiratory dysfunction or failure including dyspnea and palpitation, and the critical type is characterized by the domination of the symptom dyspnea.

Our data showed the AFAPC of symptoms did not continue to increase with the aggravation of the disease before the patients transition from the moderate type into severe type and from the severe type into critical type. However, the AFPC on the first day after transition into the severer type was significantly higher than that on the day of the transition but decreased thereafter. The reduction of AFAPC before the transitions was related to the gradual reduction of virus-induced upper respiratory tract associated symptoms. The reduction of symptoms after the transitions was related to the aggravation of consciousness disorder, the use of auxiliary ventilation and the dyspnea. These results suggest that reduction of symptoms does not represent the decreased severity of COVID-19 for the severe and critical cases. Specific symptoms related to respiratory functions and CT should be applied in the diagnose of severity.

Our classification in symptoms indicated that the COVID-19 symptoms mainly belonged to the symptoms related to respiratory tract infection (with a proportion of frequency 25.8%), respiratory function damage (48.1%), and digestive system (23.6%). In addition, two cardiovascular symptoms including palpitation and chest distress with a collective proportion of frequency 0.94% were included into the symptoms associated with respiratory function damage. These results were consistent with the published studies ^[12, 26-28]^. The proportion of other systems was 2.2%. Therefore, the COVID-19 is characterized by pathophysiological damages mainly in the respiratory, digestive and cardiovascular systems. In this result, we classified the poor appetite into digestive system symptom, but this symptom usually regarded associated with various system and/or factors, which might influenced the result a little.

We identified a total of 17 major symptoms. Our data showed that symptoms including fever, chills, nasal obstruction, runny nose which were related to upper respiratory tract infection disappeared around the day 15 after onset. The time of disappearance was similar to that of common virus infection ^[29, 30]^.The digestive system symptoms nausea, vomiting, and diarrhea, and palpitation and fatigue disappeared from the day 15 to day 30 after onset, earlier than respiratory system symptoms, suggesting that recovery from heart and digestive system damages was earlier than that from the respiratory damage, and the respiratory system function recovered very slowly. The remaining symptoms 30 days after symptom onset mainly were respiratory disorders, and subsequent respiratory insufficiency.

The major symptoms can be classified into two categories according to the changes of their frequencies. The first includes fever, poor appetite, cough, expectoration, shortness of breath, chills, pharyngeal discomfort, myalgia. Their frequencies are the highest right since onset and decreased gradually afterwards. The second includes fatigue, nausea, vomiting, diarrhea, abdominal discomfort, dizziness, dyspnea and palpitation. Their frequencies are increased since onset of COVID-19 and then decreased after reaching peak values. Interestingly, the first category of symptoms belongs to the systemic and local tissue damage and inflammatory response associated symptoms ^[31]^, mainly systemic and respiratory symptoms. The second category of symptoms belongs to respiratory and digestive organ dysfunction symptoms such as nausea, vomiting, diarrhea and abdominal discomfort ^[32-34]^. These results support that symptoms related with respiratory system, digestive system and cardiovascular systems are the main symptoms of COVID-19, and they are the main targets of COVID-19. This is consistent with the findings of pathological anatomy ^[1, 35, 36]^.

## Conclusion

The symptoms of COVID-19 cases mainly related to upper respiratory tract infection, cardiopulmonary function, and digestive system. The symptoms in the mild type and the early stage are mainly related to upper respiratory tract infection, and all other stages are characterized mainly by the cardiopulmonary function related symptoms and digestive system symptoms. Dyspnea was a marker for the critical type and dyspnea, shortness of breath, and chest distress were associated with death. The symptoms related to breathing are the characteristic symptoms of COVID-19. A total of 17 symptoms have been identified to be major symptoms of COVID-19, including fever, poor appetite, cough, expectoration, shortness of breath, chest distress, fatigue, diarrhea, dyspnea, nausea, chills, abdominal discomfort, vomiting, pharyngeal discomfort, palpitation, dizziness, and myalgia.

## Data Availability

Anyone who wishes to obtain the original data of this study with reasonable purposes can contact the correspondent author via email.

## References

1. Lamers MM, Beumer J, van der Vaart J, et al. SARS-CoV-2 productively infects human gut enterocytes. Science. 2020; 369: 50–54.

2. Wang QH, Zhang YF, Wu LL, Niu S, et al. Structural and Functional Basis of SARS-CoV-2 Entry by Using Human ACE2. Cell. 2020;181:894–904.

3. Yan RH, Zhang YY, Li YN, et al. Structural basis for the recognition of SARS-CoV-2 by full-length human ACE2. Science. 2020;367:1444–1448.

4. Shang J, Ye G, Shi K, et al. Structural basis of receptor recognition by SARS-CoV-2. 2020;581:221–224.

5. Sica A, Vitiello P, Caccavale S, et al. Primary cutaneous DLBCL non-GCB type: challenges of a rare case. Open Med (Wars). 2020;19:119–125.

6. World Health Organization: Coronavirus disease 2019 (COVID-19) situation report 2020. https://covid19.who.int/.

7. Kroenke K. Studying symptoms: sampling and measurement issues. Ann Intern Med. 2001;1:844–853.

8. Zhou F, Yu T, Du RH, et al. Clinical course and risk factors for mortality of adult inpatients with COVID-19 in Wuhan, China: a retrospective cohort study. Lancet. 2020;395:1054–1062.

9. Chen HJ, Guo JJ, Wang C, et al. Clinical characteristics and intrauterine vertical transmission potential of COVID-19 infection in nine pregnant women: a retrospective review of medical records. Lance. 2020;395:809–815.

10. Helms J, Kremer S, Merdji H, et al. Neurologic Features in Severe SARS-CoV-2 Infection. N Engl J Med. 2020;382:2268–2270.

11. Fried JA, Ramasubbu K, Bhatt R, et al. The Variety of Cardiovascular Presentations of COVID-19. Circulation. 2020;141:1930–1936.

12. Zheng YY, Ma YT, Zhang JY, et al. COVID-19 and the cardiovascular system. Nature reviews Cardiology. 2020;17:259–260.

13. Clerkin KJ, Fried JA, Raikhelkar J, et al. COronavirus disease 2019 (COVID-19) and cardiovascular disease. Circulation. 2020;140:1648–1655.

14. Lin W, Hu LF, Zhang Y, et al. Single-cell Analysis of ACE2 Expression in Human Kidneys and Bladders Reveals a Potential Route of 2019-nCoV Infection. 2020; https://www.biorxiv.org/content/10.1101/2020.02.08.939892v1.

15. Diaz-Guimaraens B, Dominguez-Santas M, Suarez-Valle A, et al Selda-Enriquez G, Bea-Ardebol S, et al. Petechial skin rash associated with severe acute respiratory syndrome coronavirus 2 infection. JAMA Dermatol. 2020; doi:10.1001/jamadermatol.2020.1741.

16. Casas CG, Catala A, Hernandez GC, et al. Classification of the cutaneous manifestations of COVID-19: a rapid prospective nationwide consensus study in Spain with 375 cases. Br J Dematol. 2020;183:71–77.

17. Spinato G, Fabbris C, Polesel J, et al. Alterations in Smell or Taste in Mildly Symptomatic Outpatients With SARS-CoV-2 Infection. JAMA. 2020;323:2089–2090.

18. Xu XW, Wu XX, Jiang XG, et al. Clinical findings in a group of patients infected with the 2019 novel coronavirus(SARS-Cov-2) outside of Wuhan, China: retrospective cases series. BMJ. 2020; doi: 10.1136/bmj.m606.

19. Young BE, Ong SWX, Kalimuddin S, et al. Epidemiologic Features and Clinical Course of Patients Infected With SARS-CoV-2 in Singapore. JAMA. 2020;323:1488–1494.

20. Wang DW, Hu B, Hu C, et al. Clinical characteristics of 138 hospitalized patients with 2019 novel coronavirus-infected pneumonia in Wuhan, China. JAMA. 2020;323:1061–1069.

21. Natinal Health Commission& National Administration of Traditional Chinese Medicine. Diagnosis and Treatment Protocol for Novel Coronavirus Pneumonia(Trial version 7). Chin Med J (Engl). 2020;133:1087–1095.

22. Weiss P, Murdoch DR. Clinical course and mortality risk of severe COVID-19. Lancet. 2020;395:1014–1015.

23. Bhatraju PK, Ghassenieh BJ, Nichols M, et al. COVID-19 in critically III patients in the seattle region-case series. N Engl J Med. 2020;382:2012–2022.

24. Chen NS, Zhou M, Dong X, et al. Epidemiological and clinical characteristics of 99 cases of 2019 novel coronavirus pneumonia in Wuhan, China: a descriptive study. Lancet. 2020;395:507–513.

25. Linder KA, Malani PN. Respiratory syncytical virus. JAMA. 2017;317:98.

26. Wolfel R, Corman VM, Guggemos W, et al. Virological assessment of hospitalized patients with COVID-2019. Nature. 2020;581:465–469.

27. Xu Z, Shi L, Wang YJ, Zhang JY, et al. Pathological findings of COVID-19 associated with acute respiratory distress synfrome. Lancet Respir Med. 2020;8:420–422.

28. Nobel YR, Phipps M, Zucker J, et al. Gastrointestinal symptoms and coronavirus disease 2019: a case-control study from the United States. Gastroenterology. 2020;159:373–375.

29. Vielle NJ, Garcia-Nicolas O, Esteves BIO, et al. The Human Upper Respiratory Tract Epithelium Is Susceptible to Flaviviruses. Front Microbiol. 2019;10:811.

30. Chung KF, Pavord ID. Prevalence, pathogenesis, and causes of chronic cough. Lancet. 2008;371:1364–74.

31. Jochems SP, Marcon F, Carniel BF, et al. Inflammation induced by influenza virus impairs human innate immune control of pneumococcus. Nat Immunol. 2018;19:1299–1308.

32. Devincenzo JP, McClure MW, Fry J. ALS-008176 for respiratory syncytial virus infection. N Engl J Med. 2016;374:1391-2/

33. Arasaratnam R. ALS-008176 respiratory syncytial virus infection. 2016;374:1391.

34. White JP, Xiong SS, Malvin NP, et al. Intestinal dysmotility syndromes following systemic infection by flaviviruses. Cell. 2018;175:1198–1212.

35. Barton LM, Duval EJ, Stroberg E, et al. COVID-19 autopsies, Oklahoma, USA. Am J Clin Pathol. 2020;153:725–733.

36. Wichmann D, Sperhake JP, Lutgehetmann M, et al. Autopsy Findings and Venous Thromboembolism in Patients With COVID-19. Ann Intern Med. 2020;173:268–277.

